# Reproducible spectral CT thermometry with liver-mimicking phantoms for image-guided thermal ablation

**DOI:** 10.1101/2023.10.04.23296423

**Authors:** Leening P. Liu, Rizza Pua, Derick N. Rosario-Berrios, Olivia F. Sandvold, Amy E. Perkins, David P. Cormode, Nadav Shapira, Michael C. Soulen, Peter B. Noël

**Author notes:** Corresponding Authors: Leening P. Liu, Peter B. Noël.

## Abstract

**Objectives:** Evaluate the reproducibility, temperature sensitivity, and radiation dose requirements of spectral CT thermometry in tissue-mimicking phantoms to establish its utility for non-invasive temperature monitoring of thermal ablations.

**Materials and Methods:** Three liver mimicking phantoms embedded with temperature sensors were individually scanned with a dual-layer spectral CT at different radiation dose levels during heating and cooling (35 to 80 °C). Physical density maps were reconstructed from spectral results using a range of reconstruction parameters. Thermal volumetric expansion was then measured at each temperature sensor every 5°C in order to establish a correlation between physical density and temperature. Linear regressions were applied based on thermal volumetric expansion for each phantom, and coefficient of variation for fit parameters was calculated to characterize reproducibility of spectral CT thermometry. Additionally, temperature sensitivity was determined to evaluate the effect of acquisition parameters, reconstruction parameters, and image denoising. The resulting minimum radiation dose to meet the clinical temperature sensitivity requirement was determined for each slice thickness, both with and without additional denoising.

**Results:** Thermal volumetric expansion was robustly replicated in all three phantoms, with a correlation coefficient variation of only 0.43%. Similarly, the coefficient of variation for the slope and intercept were 9.6% and 0.08%, respectively, indicating reproducibility of the spectral CT thermometry. Temperature sensitivity ranged from 2 to 23 °C, decreasing with increased radiation dose, slice thickness, and iterative reconstruction level. To meet the clinical requirement for temperature sensitivity, the minimum required radiation dose ranged from 20, 30, and 57 mGy for slice thickness of 2, 3, and 5 mm, respectively, but was reduced to 2 mGy with additional denoising.

**Conclusions:** Spectral CT thermometry demonstrated reproducibility across three liver-mimicking phantoms and illustrated the clinical requirement for temperature sensitivity can be met for different slice thicknesses. Moreover, additional denoising enables the use of more clinically relevant radiation doses, facilitating the clinical translation of spectral CT thermometry. The reproducibility and temperature accuracy of spectral CT thermometry enable its clinical application for non-invasive temperature monitoring of thermal ablation.

## Introduction

Thermal ablation has become increasingly utilized for minimally invasive treatment of tumors in patients with low disease burdens as an alternative to surgical resection^1,2^. In each of the different methods of thermal ablation (radiofrequency, microwave, laser, high intensity focused ultrasound), an ablation applicator focally heats tumor tissue and a surrounding safety margin to a lethal threshold of 60 °C that induces cell death within seconds^3,4^. Even with improvements in ablation technology, local tumor recurrence rates remain undesirably high, ranging from 2 to 17% for microwave ablation^5–10^ compared to 9 to 37% for radiofrequency ablation, an older technology^7,10–14^. These local recurrences are strongly associated with an insufficient coverage of the tumor and the ablative safety margin around the tumor^15–17^. Both can be assessed with visual/cognitive registration of the tumor on pre-ablation unenhanced CT and the estimated ablation zone on contrast-enhanced CT acquired immediately post-ablation^18,19^. With the addition of ablation confirmation software, automated registration has helped to identify tumors with insufficient margins, thus spurring additional treatment and increasing the percent of treated tumors with a sufficient margin from 61% to 77%^20^. Despite this increased effectiveness, necrotic tissue on contrast-enhanced CT immediately post-ablation is not only hard to distinguish from tumor tissue^21^ but continues to expand at least 24 hours after the procedure, thus is not predictive of total cell death^22^. Comparatively, using intra-tissue temperature as a predictor offers strong guidance for delineating the ablation zone. Furthermore, it can be monitored in real-time during the procedure to provide feedback to the interventional radiologist, potentially increasing the success of the ablation, reducing the risk of local tumor recurrence, and decreasing the risk of non-target injury to adjacent vulnerable structures, such as the pancreas and bowel.

Intraprocedural monitoring of temperature requires the generation of differential temperature maps to determine whether tissue temperature reaches and surpasses the lethal threshold. These temperature maps can be generated from conventional CT images through CT thermometry. CT thermometry simplifies thermal volumetric expansion to establish a linear, quadratic, or cubic relationship between changes in temperature and HU^23,24^. In the past few decades, CT thermometry has been evaluated in *ex vivo* tissues with different heating methods and CT scanners with a range of temperature sensitivities from −2.0 to −0.23 HU/C^23–27^. The variability in tissue characteristics and scanner performance, which leads to a wide range in temperature sensitivity, coupled with the necessity for high radiation doses, has hindered the clinical implementation of this method. Advancements of CT, specifically spectral CT, have enabled novel and improved methodologies that utilize spectral results, which are less dependent on scanner technology and patient habitus^28–30^. With dual-energy CT, the first realization of spectral CT, we previously developed and validated spectral CT thermometry^31^. It uses spectral results to calculate physical density maps that relate to temperature through thermal volumetric expansion^31^. Compared to traditional methods that rely on HU, which can reflect both changes in temperature and changes in tissue composition, the use of physical density maps isolates changes in temperature, ensuring more accurate temperature mapping^31^. The resulting strong correlation to thermal volumetric expansion demonstrated the feasibility of spectral CT thermometry and highlighted its potential utility for non-invasive temperature monitoring.

In order to progress towards *in vivo* evaluation and clinical translation, open questions about reproducibility and clinical requirements of spectral CT thermometry must be addressed. With the large range in model parameters in previously published studies of CT thermometry, reproducibility of the model with the same tissue/object is especially important to establish confidence in the method^25^. Similarly, temperature accuracy of <3 °C and a spatial resolution of <2 mm are required to ensure that detected temperatures at a specific voxel correspond to true tissue temperatures, and consequently ablation success^32^. In particular, the temperature sensitivity requirement is uniquely intertwined with radiation dose because of noise. Noise in physical density maps decreases with radiation dose, thus improving temperature sensitivity crucial for temperature map accuracy. Specifically, tissue heterogeneity in our previous study masked physical density noise^31^, suggesting the need for reproducible and homogeneous phantoms for evaluation of this tradeoff as well as reproducibility of the model.

This study aimed to assess the reproducibility of spectral CT thermometry with liver-mimicking gel phantoms. We also examined the effect of radiation dose and reconstruction parameters on temperature sensitivity, both at baseline and with additional denoising, and determined the corresponding radiation dose required to achieve acceptable temperature sensitivity. Our results demonstrated strong reproducibility and sufficiently low temperature sensitivity and radiation doses, supporting the clinical translation of spectral CT thermometry for non-invasive temperature monitoring during thermal ablations.

## Materials and Methods

### Phantom synthesis and characterization

To evaluate reproducibility of spectral CT thermometry and its temperature sensitivity for liver ablation via thermal volumetric expansion, a homogeneous tissue-mimicking phantom as described in Negussie et al. was modified to mimic the attenuation of liver tissue^33^. Briefly, 1 g of ammonium persulfate (Sigma Aldrich, St. Louis, Missouri) was dissolved in 2 mL of deionized water and then vortexed and sonicated until homogeneous. Separately, 290 mL of degassed and deionized water and 200 mL of 40% acrylamide/bisacrylamide (Sigma Aldrich, St. Louis, Missouri) were mixed by magnetic stirring. The ammonium persulfate solution and 1 mL of N,N,N’,N’-tetramethylethylenediamine (Sigma Aldrich, St. Louis, Missouri) were then added in quick succession to start and catalyze polymerization, respectively. The solution was then placed in a tightly sealed plastic container and placed at 4 °C for an hour. The phantom was then moved to room temperature to continue to solidify until use.

While mass density, tissue conductivity, and thermal diffusivity of the original phantom were similar to that of human soft tissues, the attenuation profile did not match liver as described by ICRU 46 Report^34^. As a result, varying amounts of calcium chloride dihydrate (Sigma Aldrich, St. Louis, Missouri) were added until the profile was adequately equivalent to the expected attenuation of liver tissue between 40 and 200 keV (± 5 HU). Each modified phantom was evaluated by scanning the phantom on a dual-layer spectral CT (IQon Spectral CT, Philips Healthcare, Eindhoven, Netherlands) at a tube voltage of 120 kVp. Full scanning and reconstruction parameters are in Table 1. Virtual monoenergetic images (VMI) were then collected every 10 keV between 40 and 200 keV, and regions of interest (ROI) were placed in the center of the phantom to measure attenuation. The resulting spectral curve was compared to the corresponding spectral curve for human liver from ICRU 46^34^. Ultimately, to match the attenuation curve, 5.8 g of calcium chloride dihydrate (Sigma Aldrich, St. Louis, Missouri) was dissolved in 10 mL of deionized water and added to the acrylamide and deionized water solution prior to polymerization. Additionally, tissue conductivity, volumetric heat capacity, and thermal diffusivity were measured with a thermal analyzer (Tempos Thermal Properties Analyzer, Meter Environment, Pullman, Washington). After confirmation of these phantom properties, the phantom was deemed liver-mimicking and sufficiently accurate for spectral CT thermometry of liver.

**Table 1.**
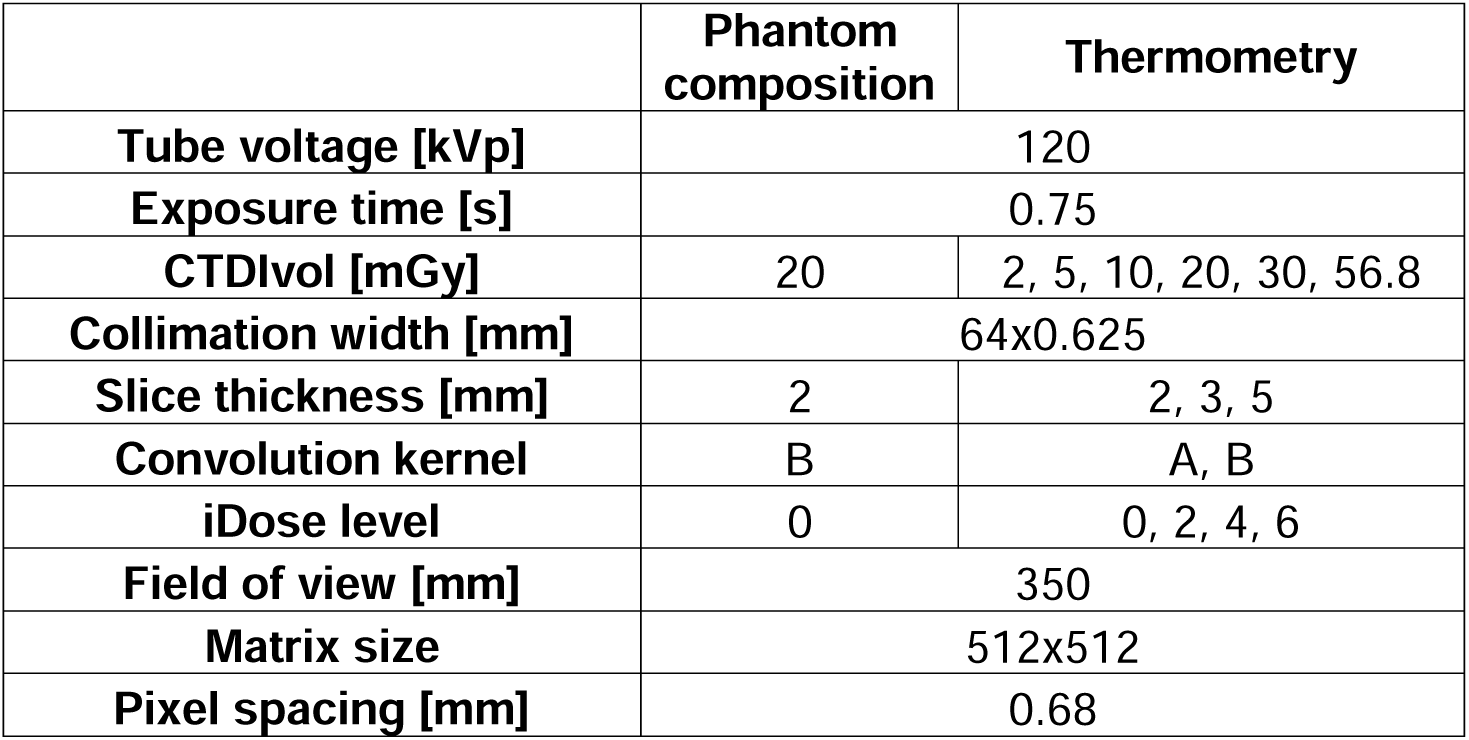
Acquisition and reconstruction parameters.

|  | **Phantom**<br>**composition** | **Thermometry** |
| --- | --- | --- |
| **Tube voltage [kVp]** | 120 |  |
| **Exposure time [s]** | 0.75 |  |
| **CTDIvol [mGy]** | 20 | 2, 5, 10, 20, 30, 56.8 |
| **Collimation width [mm]** | 64x0.625 |  |
| **Slice thickness [mm]** | 2 | 2, 3, 5 |
| **Convolution kernel** | B | A, B |
| **iDose level** | 0 | 0, 2, 4, 6 |
| **Field of view [mm]** | 350 |  |
| **Matrix size** | 512x512 |  |
| **Pixel spacing [mm]** | 0.68 |  |

For thermometry experiments, a phantom was prepared two to three days prior to each of the three repetitions. Before phantom synthesis, a 3D printed guide was glued to the lid of the plastic container to help guide two temperature probes for ground truth measurements. A fiber Bragg grating (FBG) temperature sensor (FiSpec FBG X100, FiSens GmbH, Braunschweig, Germany) was mounted 2 cm from the center and contains four sensors along its length, while an optical fiber sensor (Fiber Optic Temperature Sensor, Omega Engineering, Norwalk, Connecticut) was placed 3 cm from the center and only measures temperature at the tip of the fiber. In total, temperature was measured at five different locations. Both fibers were inserted into the phantom immediately after polymerization was initialized to ensure that they were aligned and embedded.

### Image acquisition

We assessed the reproducibility and temperature sensitivity of spectral CT thermometry using three phantoms subjected to a range of temperatures in a water bath. Each liver-mimicking phantom was placed in a plastic bucket with an immersion heater (Heet-O-Matic Immersion Heater, Ulanet, Bristol, Connecticut) for continuously heating the water and two thermocouples (Type K Thermocouple, Pico Technology, St. Neots, United Kingdom) for monitoring the temperature of the water next to and across from the heater (Figure 1). Boiled water was carefully poured into the bucket to completely immerse the phantom. After water temperatures cooled to 60 °C, the immersion heater was turned on to further heat the water until the phantom reached approximately 80 °C. Water temperatures rose on average 3.5 °C per minute. Then, ice was added to the water approximately every 20 minutes until the phantom cooled down to 35 °C (Figure 2). During heating and cooling phases, the phantom was scanned with dual-layer spectral CT at a tube voltage of 120 kVp with axial scans and 4 cm collimation. Scans were performed approximately every 30 seconds, altering between different radiation doses at volumetric CT dose index (CTDI_vol_) of 2, 5, 10, 20, 30, and 56.8 mGy. Other acquisition parameters can be found in Table 1. The process of heating, cooling, and scanning was repeated for each of the three separate phantoms to evaluate reproducibility.

**Figure 1.**
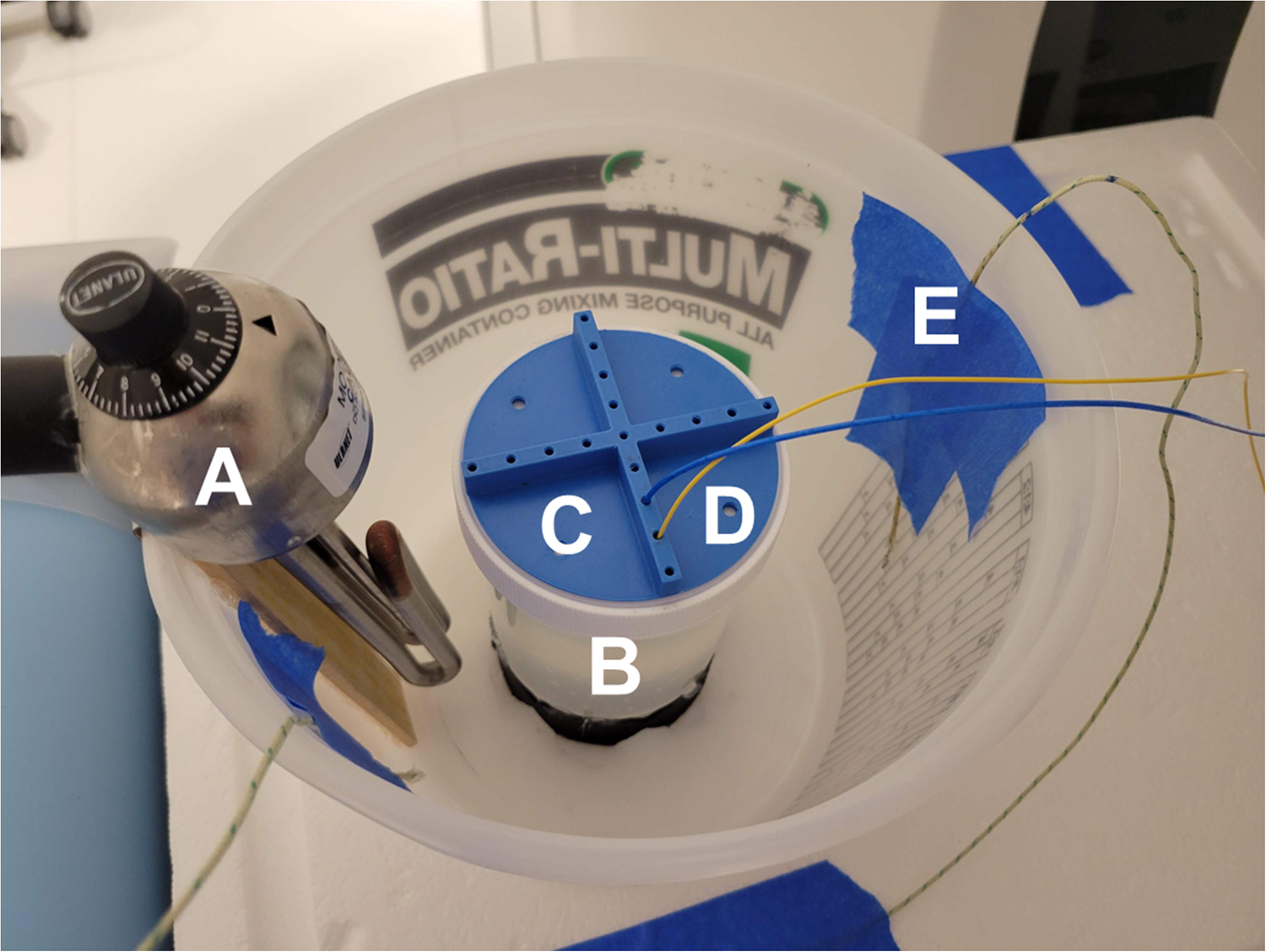
Experimental setup for reproducibility of spectral CT thermometry. Hot water and an immersion heater (A) were utilized to heat a liver-mimicking phantom (B). A 3D printed guide (C) helped accurately place thermometers (D), a FBG grating fiber and optical fiber thermometer, to measure internal phantom temperatures. Additional thermocouples (E) were placed in the water to monitor temperatures.

**Figure 2.**
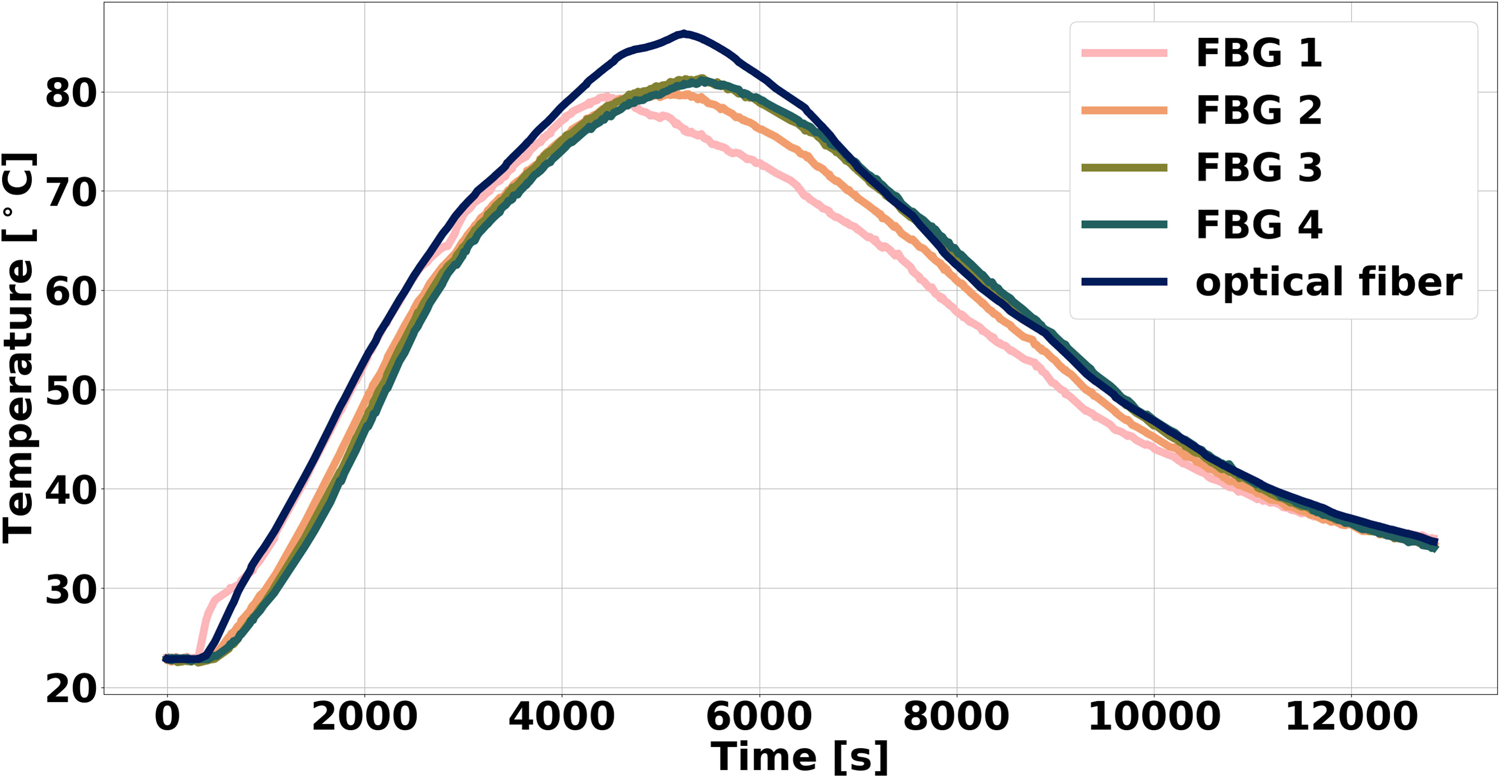
Measured internal phantom temperature during heating and cooling of phantom. Rise in temperatures occurred with heating from both hot water and an immersion heater, while cooling resulted from addition of ice into the water bath.

Physical density maps were utilized to relate physical density and temperature through thermal volumetric expansion. First, spectral results, specifically VMI at 70 keV and effective atomic number maps, were generated for acquisitions between approximately 35 and 80 °C at an interval of 5 °C for each radiation dose. Reconstructions varied in slice thickness (2, 3, 5 mm), iterative reconstruction (IR) level (iDose^4^ level 0, 2, 4, 6), and reconstruction kernel (A, B) to investigate the effect of parameters on temperature sensitivity. Using VMI 70 keV and effective atomic number maps, physical density maps were calculated by applying a previously developed model^31^. Then, in order to maximally decrease noise and thus decrease the minimum required dose for clinical translation, additional denoising with non-local means was also applied to spectral results prior to generating physical density maps. Non-local means denoising uses a pixel-weighted average based on the similarity to the region^35^. From the reconstructed spectral results, physical density maps were generated by using a previously developed spectral physical density model. On these physical density maps, the tip of the fiber optic temperature sensor, the tip of the FBG fiber, and the angle of the FBG fiber were manually determined. With this information, locations of the five temperature sensors were calculated. ROIs were then placed opposite of the temperature sensors and equidistant to the center axis of the phantom to avoid the attenuation of the fibers (Figure 3). Placement of these ROIs were possible because of the homogenous nature of the phantom and the left-right symmetry of the heating of the phantom. Mean and standard deviation of physical density at each temperature sensor were then recorded at each corresponding temperature and reconstruction parameter combination. Additional denoising and all analysis was performed with Python.

**Figure 3.**
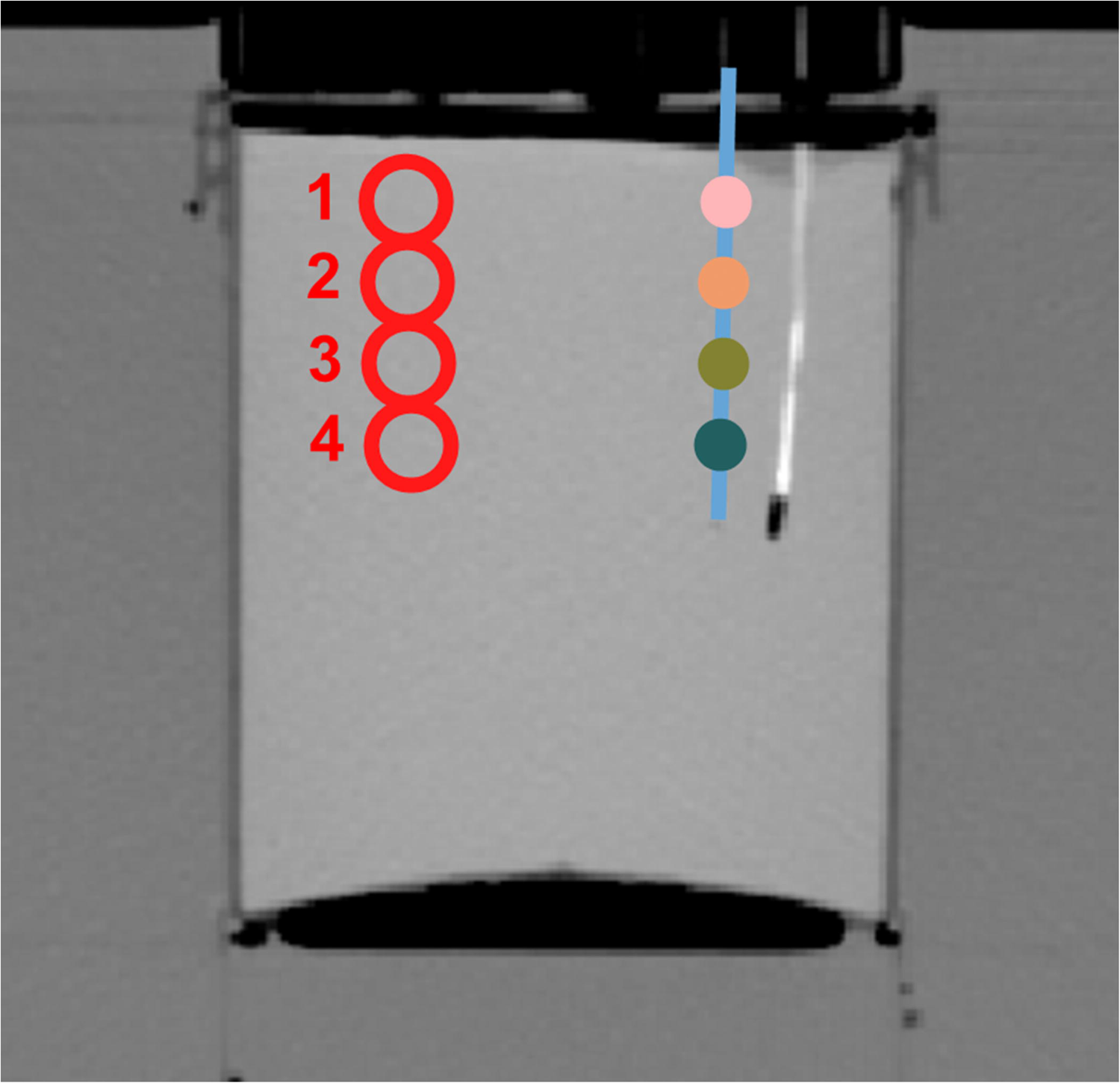
Corresponding physical density measurement for each FBG along the fiber. The high attenuating fiber corresponds to the optical fiber with a single sensor at its end while the blue line represents the FBG fiber with its sensors along its length (dots). ROIs (red circles) were placed symmetrically from each temperature sensor to avoid including thermometers in ROIs.

### Spectral CT reproducibility

For each of the three phantoms, thermal volumetric expansion was utilized to relate physical density to change in temperature:

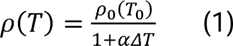

where *ρ* is physical density at temperature *T*, *ρ*_0_ is the initial physical density at the initial temperature *T*_0_, *ΔT* is the difference in temperature, and *α* is the thermal volumetric expansion constant. Specifically, a linear regression was applied to the relationship between the inverse of the physical density and change in temperature for an IR level 0, reconstruction filter A, and slice thickness of 3 mm.

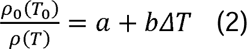

The initial physical density and temperature were calculated as the mean physical density and temperature across radiation dose at approximately 35 °C. Corresponding slope, intercept, and Pearson’s correlation coefficient (R) were recorded for each temperature sensor and repetition. Mean and standard deviation of the model parameters were calculated. Variation of fit parameters was characterized by the resulting coefficient of variation (CV), or standard deviation divided by the mean. Additionally, scatter plots were used to illustrate the recapitulation of thermal volumetric expansion for each of the three repetitions.

### Temperature sensitivity

Temperature sensitivity, defined as the maximum error between computed and measured temperature, was determined for each combination of radiation doses and reconstruction parameters at the lowest and highest temperatures. By applying thermal volumetric expansion (Eqn. 1), temperature sensitivity was calculated through error propagation:

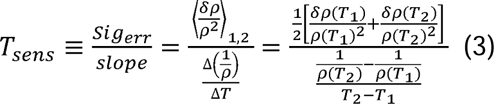

Where *sig_err_* is the propagated error of the model, *δρ* is the slope of the thermometry model, and is noise (standard deviation in an ROI) in physical density maps. As a result, temperature sensitivity is greatly dependent on noise, which can be modulated with acquisition parameters, reconstruction parameters, and denoising algorithms. To first establish baseline temperature sensitivity without external denoising, temperature sensitivity was calculated for different acquisition and reconstruction parameters. These values were then represented as a heatmap, where each subsection corresponded to a radiation dose and slice thickness combination. Also, for each slice thickness, the minimum radiation dose required to achieve clinically relevant temperature sensitivities was recorded. The same calculations were performed to illustrate the radiation dose reduction with additional post-processing^36^.

## Results

### Phantom characterization

The phantom was adequately modified to match the attenuation profile of human liver (Figure 4). While the original phantom resulted in errors as great as 15 HU for different VMIs from 40 to 200 keV, the modified liver-mimicking phantom exhibited differences in attenuation to human liver tissue ranging of only up to 4 HU. Additionally, the thermal conductivity, thermal diffusivity, and volumetric heat capacity were 0.5209 W K^-1^ m^-1^, 0.18 mm^2^/s, and 2.902 J m^-3^ K^-1^, respectively. These thermal properties corresponded to the thermal properties of human liver^37^.

**Figure 4.**
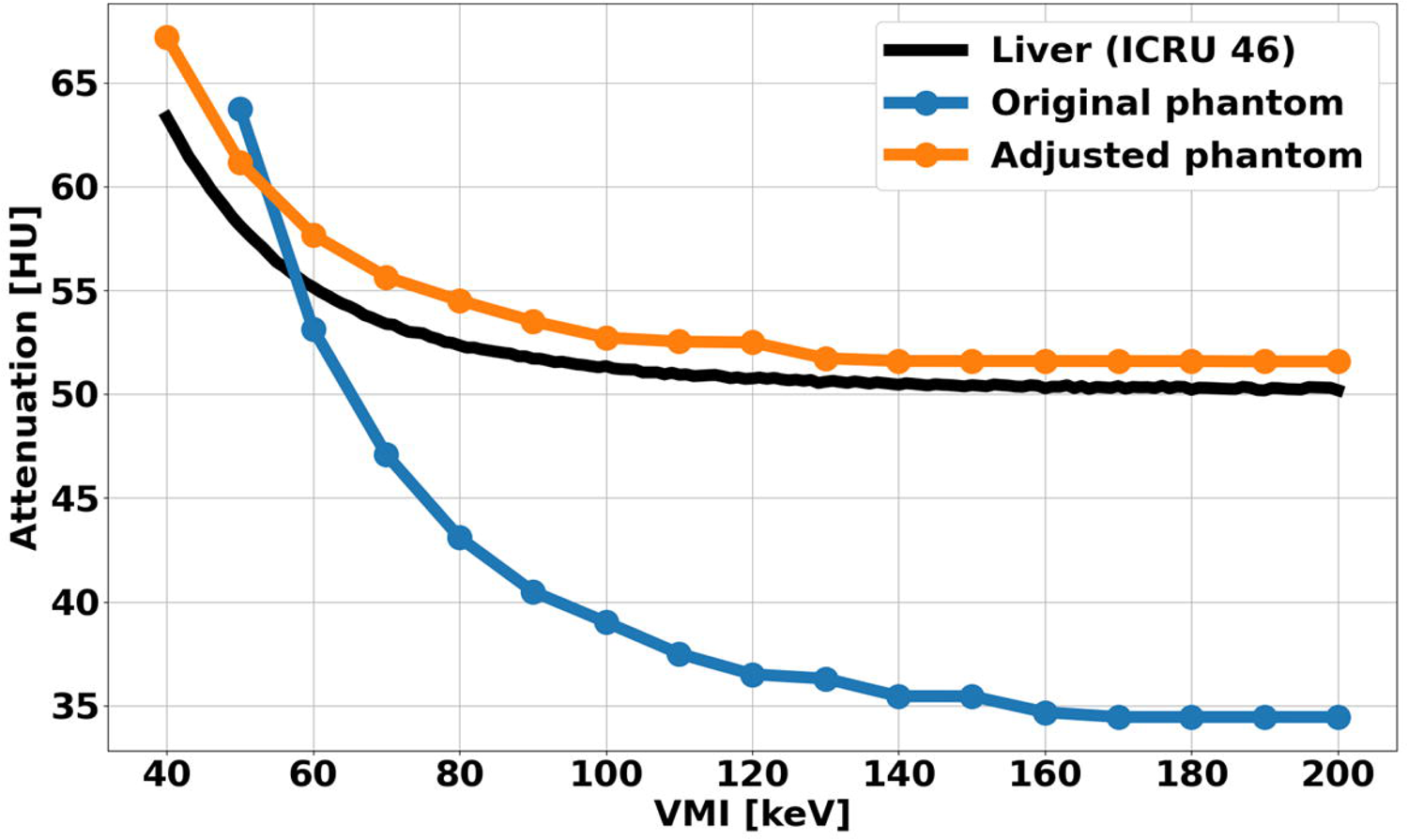
Attenuation of liver-mimicking phantom across VMI energies. Attenuation of the adjusted phantom better matched the attenuation of liver as described by ICRU 46 with a maximum difference of 4 HU.

### Spectral CT reproducibility

Liver mimicking phantoms reflected a strong relationship between the inverse of the physical density and temperature that recapitulated thermal volumetric expansion for each repetition (Figure 5). Slope and intercept measured 5.3 ± 0.5 x 10^-4^ °C^-1^ and 0.9995 ± 0.0008, respectively, across different repetitions, temperature sensors, and radiation doses without additional denoising. These values corresponded to a CV of 9.6% and 0.08% for slope and intercept, respectively, highlighting the reproducibility of spectral CT thermometry. The strong relationship was particularly demonstrated with an R of 0.989 ± 0.005 and a CV of 0.43%.

**Figure 5.**
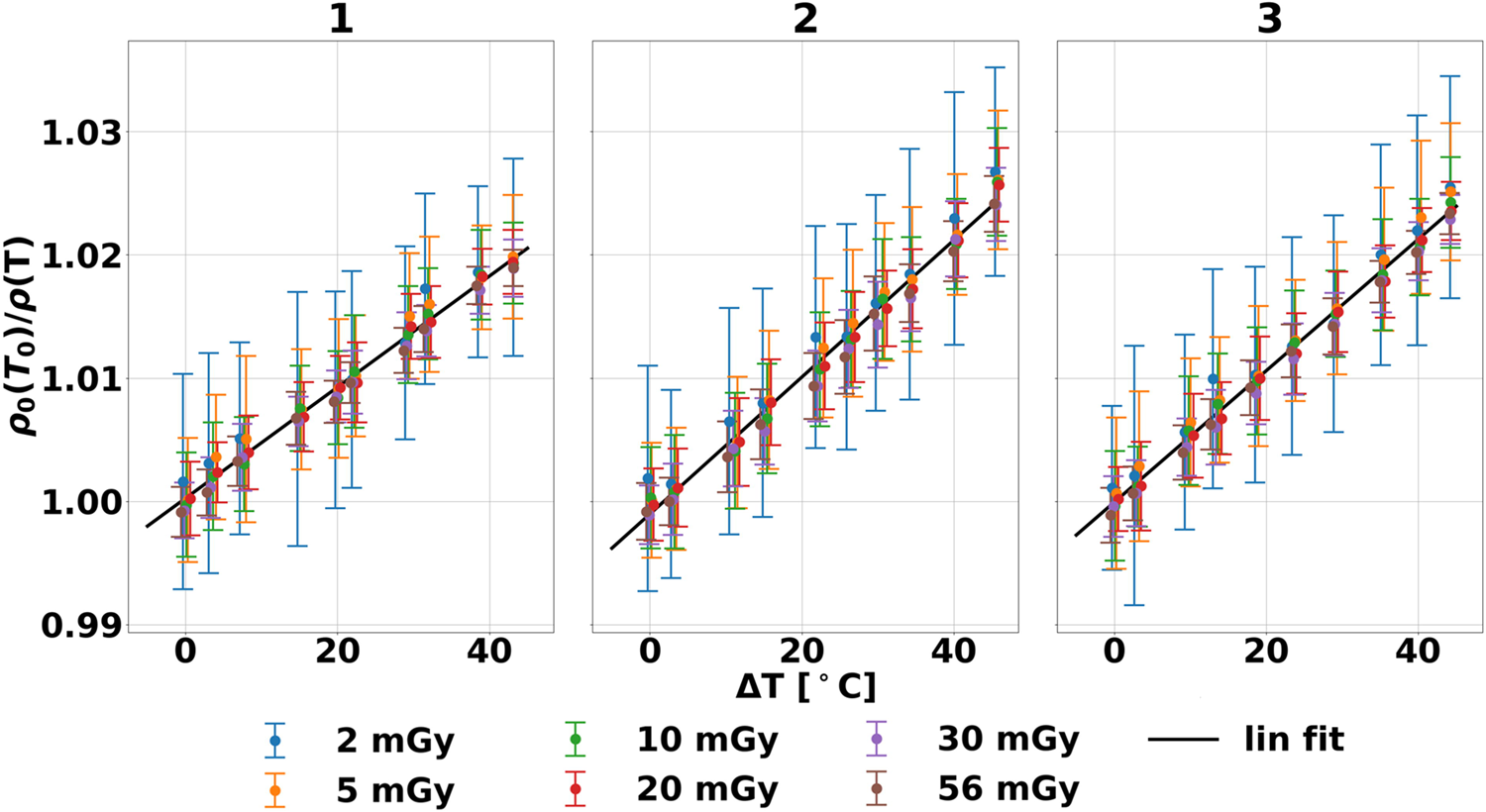
Reproducibility of spectral CT thermometry across three phantoms. The relationship between physical density and temperatures in three experiments exhibited linearity that corresponds to thermal volumetric expansion that is shown to be independent of dose. Additionally, similar behavior between the phantoms (numbered) highlighted the reproducibility of spectral CT thermometry.

### Temperature sensitivity

Temperature sensitivity decreased with increased slice thickness, iterative reconstruction level, and radiation dose for each temperature sensor (Figure 6A). They ranged from 23 to 2.2 °C, 18 to 1.9 °C, and 14 to 1.8 °C for a slice thickness of 2, 3, and 5 mm, respectively. Specifically, at 2 mGy and 2 mm slice thickness, temperature sensitivity spanned from 23 to 9.7 °C. With additional post-processing denoising, noise in physical density images decreased, and consequently, temperature sensitivity at the same parameters decreased to 1.8 to 6.8 °C (Figure 6B). As a result, the minimum radiation dose to meet the required temperature sensitivity decreased to 2 mGy for each slice thickness compared to 57, 30, and 20 mGy without denoising at slice thicknesses of 2, 3, and 5 mm, respectively.

**Figure 6.**
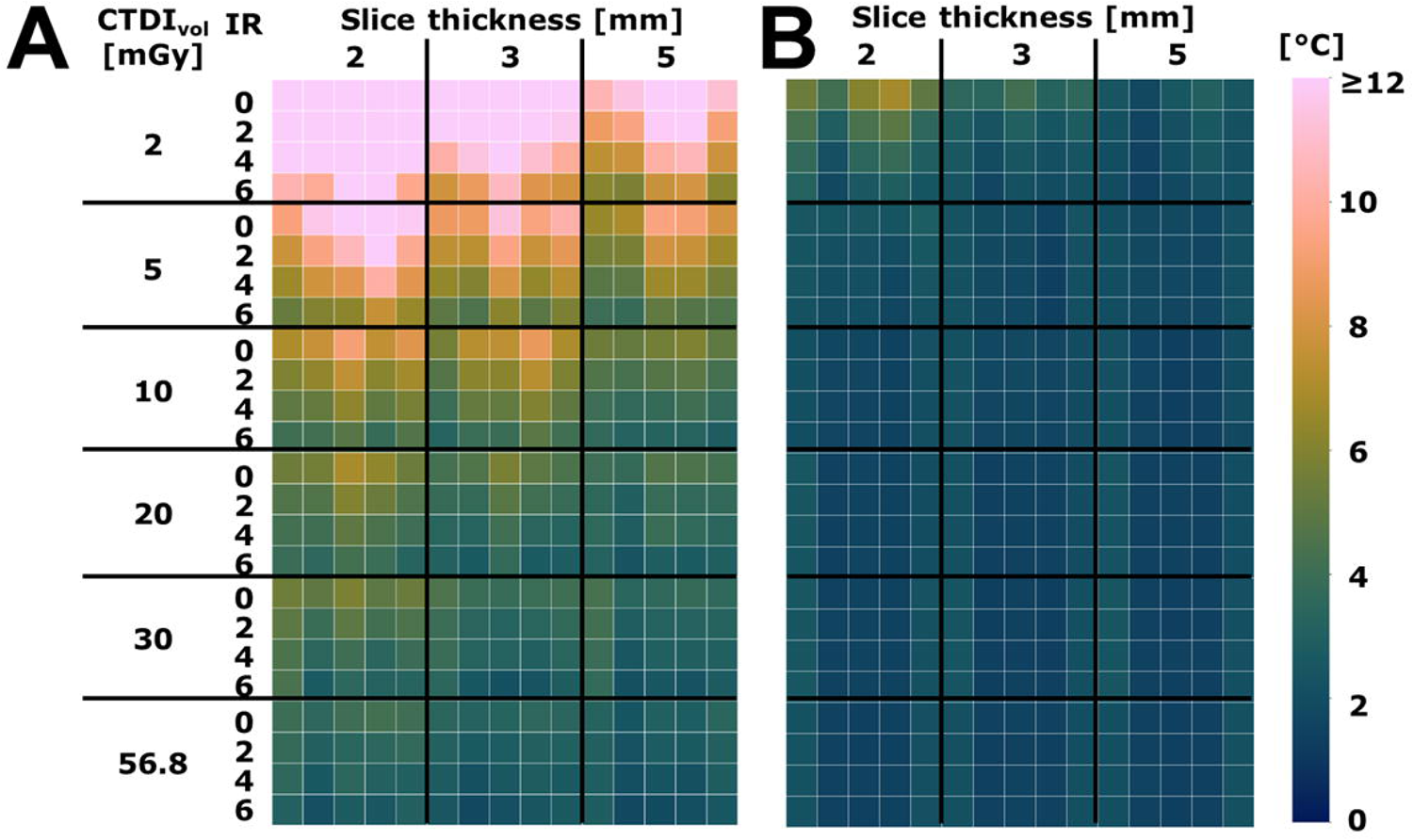
Temperature sensitivity of spectral CT thermometry. In each subsection, the columns from left to right correspond to the different temperature sensors of the FBG and optical fiber. Temperature sensitivity demonstrated a strong relationship with noise, improving with radiation dose, slice thickness, and IR level (A). Additional denoising further reduced temperature sensitivity and reduced the effect of radiation dose and reconstruction parameters (B).

## Discussion

Spectral CT thermometry demonstrated reproducibility in liver-mimicking phantoms and achieved clinically relevant temperature sensitivity by adjusting radiation dose and slice thickness. The incorporation of denoising further reduced the minimum radiation dose needed to meet clinical temperature sensitivity requirements. These findings, including reproducibility, low temperature sensitivity, and reduced radiation dose, support the clinical application of spectral CT for non-invasive temperature monitoring during liver tumor ablations.

Compared to conventional CT thermometry studies, this study not only utilized a direct model of thermal volumetric expansion but also implemented liver-mimicking phantoms, investigated the effect of the scan protocol on temperature sensitivity, and applied post-processing to address the clinical requirements of spectral CT thermometry. Liver-mimicking phantoms were utilized for the assessment of reproducibility and temperature sensitivity of spectral CT thermometry. While previous studies have used *ex vivo* and *in vivo* tissues of varying types^38–40^, these tissues are naturally heterogeneous and vary from sample to sample, both in terms of attenuation and tissue conductivity properties. Intra- and inter-sample variation of *ex vivo* tissue may thus hinder any evaluation of reproducibility as well as mask the effect of acquisition parameters on temperature sensitivity by mixing heterogeneity with image noise^31^. Liver-mimicking phantoms, on the other hand, are homogeneous and its synthesis repeatable to ensure it maintains the same properties from batch to batch. Even so, there may be some variation that then translates to the CV of the slope, likely due to the use of three separate phantoms. Other potential sources of variation for CV include the lack of additional denoising, air bubbles at the tip of temperature probes, and the similar but inconsistent heating method between phantoms. However, the CV may reflect expected variation that results from *ex vivo* and *in vivo* tissues. Future studies prior to clinical translation will require *in vivo* evaluation that accounts for both tissue heterogeneity and the heat-sink effect.

In addition to liver-mimicking phantoms, evaluation of spectral CT thermometry required a new methodology for calculating temperature sensitivity specific to the model. In conventional CT thermometry, temperature sensitivity was determined by model parameters and has ranged from −2.0 to −0.34 HU/°C^23,25,26,41^. This can be extrapolated to a temperature accuracy of 3 – 5 °C^42^. Because spectral CT thermometry implements a different model, temperature sensitivity can no longer be derived from model parameters. Instead, temperature sensitivity is derived from error propagation of thermal volumetric expansion. This methodology accounts for our recently introduced model as well as factors in the effect of noise present in CT images that may affect the accuracy of generated temperature maps. As a result, temperature sensitivity in spectral CT thermometry is acquisition and reconstruction parameter dependent. To match the accuracy of previous thermometry studies, radiation doses between 10 and 30 mGy were required for slice thicknesses ranging from 2 to 5 mm. For even lower temperature sensitivity, higher radiation doses were needed but were not optimal for clinical translation.

In order to reduce the radiation dose, additional denoising was implemented to ensure the clinical requirement for temperature sensitivity was met. While the denoising here was performed on a piecewise constant phantom^33^ and thus may not be practical for clinical translation, it provides an example of post processing that may significantly benefit the clinical translation of spectral CT thermometry. Another example, though not required nor utilized in this study, is metal artifact reduction for ablation applicators. These metal artifacts reduce image quality and quantitative integrity^43^ that would propagate to temperature maps, particularly in the vicinity of the applicator itself. As a result, quantitatively accurate metal artifact reduction is required for accurate and clinically useful non-invasive temperature monitoring. Such development and evaluation of metal artifact reduction algorithms for spectral CT thermometry are currently in progress and will ensure artifact free and reliable temperature maps.

Reproducibility and clinically relevant temperature sensitivity of spectral CT thermometry bolster its utility for non-invasive temperature monitoring to provide real-time feedback to interventional radiologists during thermal ablation. Unlike spectral CT thermometry, current methods require interventional radiologists to extrapolate between pre-ablation and post-ablation contrast-enhanced CT scans (visual/cognitive registration) to establish a complete ablation and sufficient minimal ablative margin^18,19^. Even with the addition of ablation confirmation software, follow-up magnetic resonance imaging still indicated that only 77% of ablations had a sufficient minimal ablative margin^20^, suggesting the need for other techniques. Specifically, spectral CT thermometry’s intraprocedural, volumetric temperature maps cannot only discern the tumor volume that has already reached the lethal threshold but also track temperatures of surrounding critical structures to minimize thermal damage. With such a tool and its information, interventional radiologists can adjust ablation duration in real-time to guarantee a complete ablation and a sufficient minimal ablative margin but requires confidence in spectral CT thermometry. Reproducibility of spectral CT thermometry builds this confidence by demonstrating thermal volumetric expansion and the same model parameters even with different phantoms. Additionally, both temperature sensitivity and radiation dose requirement are met by adjusting acquisition parameters and applying additional post-processing. Each of these three aspects enhance spectral CT thermometry’s utility for non-invasive temperature monitoring in clinical thermal ablation procedures.

In conclusion, physical density maps demonstrated a strong, reproducible relationship with temperature in liver-mimicking phantoms with temperature sensitivities that varied with acquisition and reconstruction parameters. Temperature sensitivity was demonstrated at clinically relevant levels and the required radiation dose was lowered to 2 mGy with denoising. By meeting the requirements for temperature sensitivity and radiation dose, spectral CT thermometry can increasingly facilitate the clinical adoption of non-invasive temperature monitoring during thermal ablation procedures, thereby reducing the risk of local tumor recurrence.

## Data Availability

All data produced in this study are available upon reasonable request.

## Acknowledgements

We acknowledge support from Philips Healthcare, National Institutes of Health (R01EB030494), and Society of Interventional Radiology Foundation (Allied Scientist Training Grant).

## Abbreviations

CT: computed tomography
HU: Hounsfield Units
VMI: virtual monoenergetic images
ROI: regions of interest
FBG: fiber Bragg grating
CTDI_vol_: volumetric CT dose index
IR: iterative reconstruction
R: Pearson’s correlation coefficient
CV: coefficient of variation

